# Multi-type branching and graph product theory of infectious disease outbreaks

**DOI:** 10.1101/2020.10.09.20210252

**Authors:** Alexei Vazquez

## Abstract

The heterogeneity of human populations is a challenge to mathematical descriptions of epidemic outbreaks. Numerical simulations are deployed to account for the many factors influencing the spreading dynamics. Yet, the results from numerical simulations are often as complicated as the reality, leaving us with a sense of confusion about how the different factors account for the simulation results. Here, using a multi-type branching together with a graph tensor product approach, I derive a single equation for the effective reproductive number of an infectious disease outbreak. Using this equation I deconvolute the impact of crowd management, targeted testing, contact heterogeneity, stratified vaccination, mask use and smartphone tracing app use. This equation can be used to gain a basic understanding of infectious disease outbreaks and their simulations.

---

Infectious diseases spread in heterogenous populations of susceptible individuals. There is variability in the number of potential contacts [1], age groups [2] and adherence to non-pharmaceutical interventions [3]. These heterogeneities may sound too complex to be handle by means of analytical descriptions, leaving us with the choice of numerical simulations. Numerical simulations are the right context to introduce all kinds of parametrizations [3–5]. Yet, we want a basic understanding as well, albeit sacrificing numerical precision. Here I demonstrate that a combination of multi-type branching process theory and graph tensor products disentangles the contributions of different factors and containment strategies to the outbreak dynamics.

The susceptible, infected and removed (SIR) model is a good representation of infectious disease outbreaks when the recovery from the disease confers immunity. In the case of COVID-19 it is not clear how long a person remains immune to the disease after infection, but it is expected to be at least of the order of months. In the SIR model the disease states are susceptible to acquire the disease, infected and removed due to death or recovery from the disease. Infected individuals can transmit the disease to susceptible individuals when they are in contact. In the case of COVID-19, contact means physical proximity for a certain amount of time. In the case of HIV contact means sexual intercourse, syringe-needle sharing or mother giving birth baby.

Some individuals visit crowded places during a day, getting in contact with several people. Others work at home and get in contact with few house mates. With relevance to sexually transmitted diseases, there is a broad distribution in the number of sexual partners of individuals across a population [6]. I will call this *contact heterogeneity*. The number of physical proximity contacts in a day, or the number of sexual partners within a year, can vary from zero to 100s and it is better represented by a probability distribution.

Individuals are also different regarding their perception of containment strategies. In the ongoing COVID-19 pandemic face masks and smartphone tracing apps are not used by all individuals. For HIV and other sexually transmitted diseases additional heterogeneities include sexual orientation, condom use, drug use, among other factors. I will call this *type heterogeneity*, where a type can be any property taking values over a discrete set of small size that can have an impact on the infectious disease dynamics. The types are characterized by their frequency in the population and the mixing patterns between individuals according to type.

Another source of variability is the disease dynamics within individuals. This dynamics could be correlated with the contact or type heterogeneities. For example, the population is stratified by age and age influences the infectious dynamics within individuals. Here I focus on the contact and type heterogeneity and assume that the disease dynamics within individuals is uncorrelated from the contact and type heterogeneity. The transmission dynamics will be characterized by the generating time, denoted by *τ*, defined as the interval from the time of infection of an individual to the time it transmit the disease to a susceptible individual. I will denote by *g*(*τ*) the probability density function of the generating time.

Here I model a population of susceptible individuals as a multi-type Markov process in the limit of large populations. I will assume that the statistical properties of individuals, including to who they transmit the disease, are dictated by their types. Using the multi-type branching process formalism, I have calculated the expected number of infected individuals of epidemic outbreaks on heterogeneous populations [7]. In a nutshell, the multi-type formalism replaces the average reproductive number, an scalar, by a matrix of reproductive numbers, making an distinction between patient zero an any other infected individual. In more detail, each individual contacts other individuals at some rate *λ*. The mixing pattern is represented by the probability *e*_*ab*_ that an individual of type *a* reaches a type *b* individual upon contact. Each contact results in disease transmission with probability *r* and the effective disease transmission rate is denoted by *β* = *λr*.

Infected individuals are removed, die to recovery, isolation or death, at a rate *γ*. Under these assumptions, the reproductive number matrix for patient zero has elements

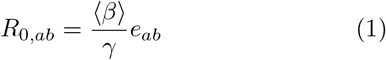

where ⟨·⟩ denotes the average over the heterogeneity of contact rates *λ* across individuals. For infected cases other than patient zero, I take into account the disease spreading bias to individuals with a higher contact rate. The patient zero can be thought as an individual selected at random from the population. Any other infected individual is not selected at random, it is found with a probability proportional to its contact rate: *β/N ⟨ β* ⟩, where *N* is the population size. Once infected, the individual found by contact will engage in new contacts at a rate *β*. Therefore, the reproductive number matrix for patients other than patient zero has elements

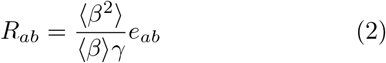

*R*_0_ gives the average number of infectious at the first generation, *R*_0_*R* at the second generation and *R*_0_*R*^*d*−1^ at the *d* generation. The actual time when an infected case at generation *d* becomes infected equals the sum of *d* generation times and it has a probability density function *g*^⋆*d*^(*t*), where the symbol ⋆ denotes convolution 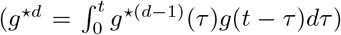. Therefore, the average number of new infected individuals at time *t* is given by (equation (36) in Ref. [7])

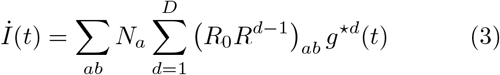

where *N*_*a*_ is the number of patients zero of type *a* and *D* is the maximum generation.

The shape of the distribution of generation times determines the functional dependence of the number of new infections with time [8]. In contrast, it does not change the epidemic threshold. Fore the sake of simplicity I will use the SIR model. For the SIR model the distribution of generating times is the distribution of recovery times. Given that recovery takes place at a constant rate, the distribution of generation times is exponential

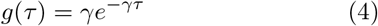

In this case, equation (3) has two limiting behaviours depending on the parameter

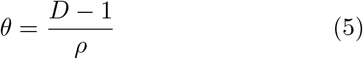

where *ρ* is the largest eigenvalue of *R* [7]. When *ρ >* 1 and *θ* ≫ 1, then for *γt* ≪ *θ* the number of new infectious grows exponentially according to

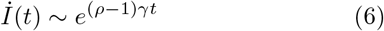

Note that *R*, and therefore *ρ*, is inversely proportional to *γ*. In contrast, when *θ* ≪ 1, then for *γt* ≫ *θ* the number of new infectious grows as a power law with an exponential cutoff

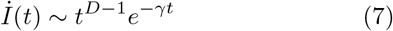

Therefore the outbreaks dynamics is determined by the largest eigenvalue of the reproductive number matrix *R* and the maximum number of generations *D*. The power low prediction has been observed in numerical simulations of virus spreading in the Internet [9] and the first COVID-19 outbreak [10].

The branching process is suitable to model the early phase of epidemic outbreaks. It has limitations to estimate the late dynamics when there is a reduction in the number of susceptible individuals. A compartment model represented by differential equations is more suitable to understand the late dynamics. Nevertheless, the key quantity of the compartment model is still the reproductive number matrix *R* [11]. Therefore I will focus on the impact of heterogeneities on the largest eigenvalue *ρ*.

The type mixing matrix, with elements *e*_*ab*_, is represented by a directed weighted graph with loops. A directed edge (arc) is drawn from *a* to *b* when *e*_*ab*_ *>* 0. Loops account for infected individuals of a given type infecting susceptible individuals of the same type. The arcs weights *e*_*ab*_ quantify the probability of finding type *a* coming from type *b*. Figure 1 illustrates type graphs associated with vaccination, mask use or smartphone use. In each case there are two types: vaccinated or not, wears mask or does not, smartphone tracing app user or not. The associated mixing matrices are 2 × 2 matrices and it is straightforward to calculate the largest eigenvalue. The challenge begins when we consider a combinations of those or other population stratifications at once. We would have to include several types and deal with matrices of largest dimension, making an analytical description cumbersome and prompting calculation errors.

**FIG. 1.**
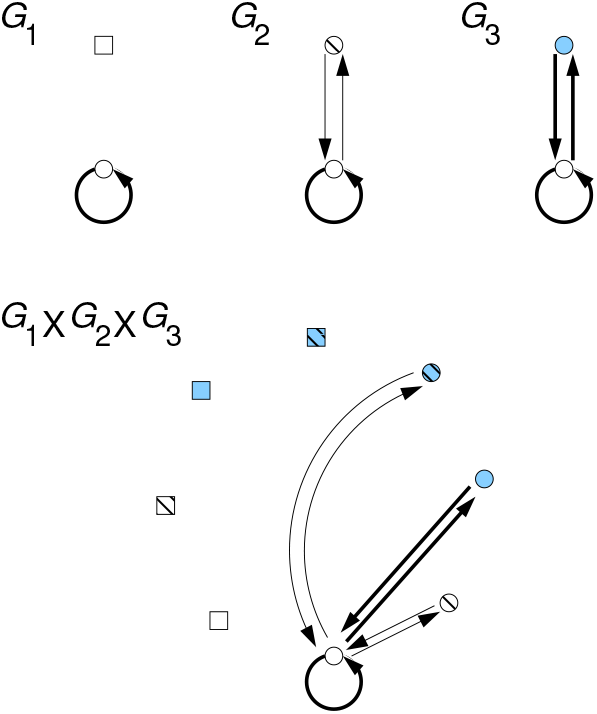
Type graphs for vaccination (*G*_1_), mask use (*G*_2_), smartphone tracing app use (*G*_3_) and their graph tensor product (*G*_1_ × *G*_2_ × *G*_3_). Open circles represent individuals not covered by the containment strategy. Squares represent vaccinated individuals, diagonal lines wearing a mask and symbol filling using the smartphone tracing app.

When different type stratifications are independent, meaning that being of one type in one stratification is uncorrelated with being of another type in another stratification, we can tackle the problem with graph tensor products. Under the assumption of independence, the type graph taking into account *n* independent population stratifications can be represented by the graph tensor product of each independent stratification

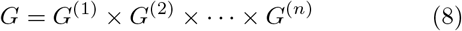

An example is shown in Fig. 1. In turn, the type mixing matrix of graph *G* can be written as a Kronecker product of the type mixing matrices of graphs *G*_*i*_,

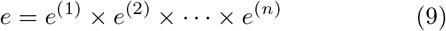

The eigenvalues of the Kronecker product of two matrixes are given by the pairwise product of the eigenvalues of each matrix (Theorem 13.12, [12]). An obvious corollary, the largest eigenvalue of *e* is equal to the product of the largest eigenvalues

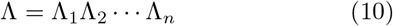

where Λ_*i*_ denotes the largest eigenvalue of *e*^(*i*)^. Finally, the largest eigenvalue of *R* in equation (2) is given by

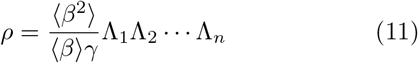

We can use equation (11) to estimate the effectiveness of mixed strategies to contain an infectious disease outbreak. To illustrate how it is done, let us consider the case of a population where crowd management, targeted testing, vaccination, mask use and smartphone tracing apps have been deployed. Crowd management reduces the contact rate *λ*, in turn reducing *β* = *λr*. The main component of targeted testing is testing the contacts traced from an infected case. Since the testing of the traced contacts effectively eliminate them from the disease transmission chain, targeted testing can be also modelled by an effective reduction of the contact rate. Therefore, crowd management and targeted testing are modelled by the transformation

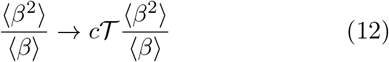

where *c* and 𝒯 are the reduction in transmission rate due to crowd management and targeted testing.

Vaccination is modelled by the type graph *G*_1_ in Fig. 1 1 and the associated type mixing matrix

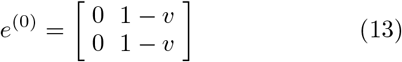

where *v* is the fraction of vaccinated individuals. More generally, we consider stratified vaccination according to a type (e.g., age group). Let *a*_*ij*_ be the mixing matrix elements of the type driving the vaccine stratification and *v*_*i*_ the fraction of *i*-type individuals that are vaccinated. When *j* is vaccinated then *e*_*ij*_ = 0. When *j* is not vaccinated it can be infected by the non-vaccinated connections and *e*_*ij*_ = *a*_*ij*_(1 −*v*_*j*_) = *a*_*ij*_(1 −*v*)*x*_*j*_, where *v* = ∑_*i*_ *v*_*i*_ and *x*_*i*_ = (1 −*v*_*i*_)*/*(1 −*v*). Once again, making use of the graph tensor product, we write the mixing matrix of stratified vaccination as

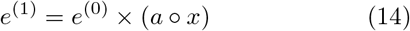

where ∘ denotes the Hadamard, element-wise, product. The largest eigenvalue of *e*^(1)^ is

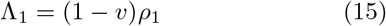

where (1−*v*) is the largest eigenvalue of *e*^(0)^ and *ρ*_1_ is the largest eigenvalue of *a ∘ x*. Note that the *x*_*i*_ can be optimized to obtain the vaccination strategy that minimizes *ρ*_1_ given a total vaccination capacity *v*.

Mask use is modelled by the type graph *G*_2_ in Fig. 1. I assume a fraction *m* of individuals wearing mask, no transmission between mask users (*e*_11_ = 0), transmission with attenuation efficiency 0 *≤ a*_1_ *<* 1 from a mask user to a non-user (*e*_12_ = *a*_1_(1 −*m*)), transmission with attenuation efficiency 0 *≤ a*_2_ *<* 1 from a non-mask user to a user (*e*_21_ = *a*_1_(1 −*m*)) and transmission between non-mask users (*e*_21_ = 1 −*m*). In this case the mixing matrix and the largest eigenvalue are

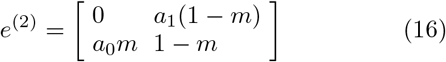

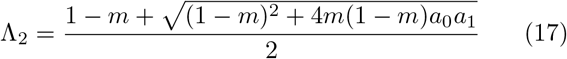

Smartphone tracing app use is modelled by the type graph *G*_3_ in Fig. 1. I assume a fraction *u* of smartphone tracing app users. The chain of transmissions between app users is truncated because of the forward and backward tracing. This will be modelled as no disease transmission between app users (*e*_11_ = 0), which is an effective approximation to be tested. With these assumptions we obtain the type mixing matrix and the largest eigenvalue

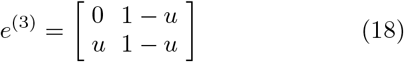

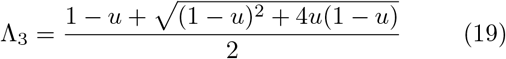

Figure (2) shows the largest eigenvalue of the different containment strategies as a function of the fraction of individuals subject to the intervention (vaccinated, mask user, smartphone tracing app user). It is evident that the largest eigenvalues associated with mask use and smartphone tracing app use are concave functions of the corresponding users fraction. Therefore, for small user fractions there is not much reduction of the largest eigenvalue. These containment strategies requires that many individuals become users. For example, 50% of mask users will reduce the reproductive number by just 20%. Furthermore, mask use is more effective that smartphone tracing app use. This is because mask use reduces the probability of transmission between mask users and nonusers, while the smartphone tracing app does not.

**FIG. 2.**
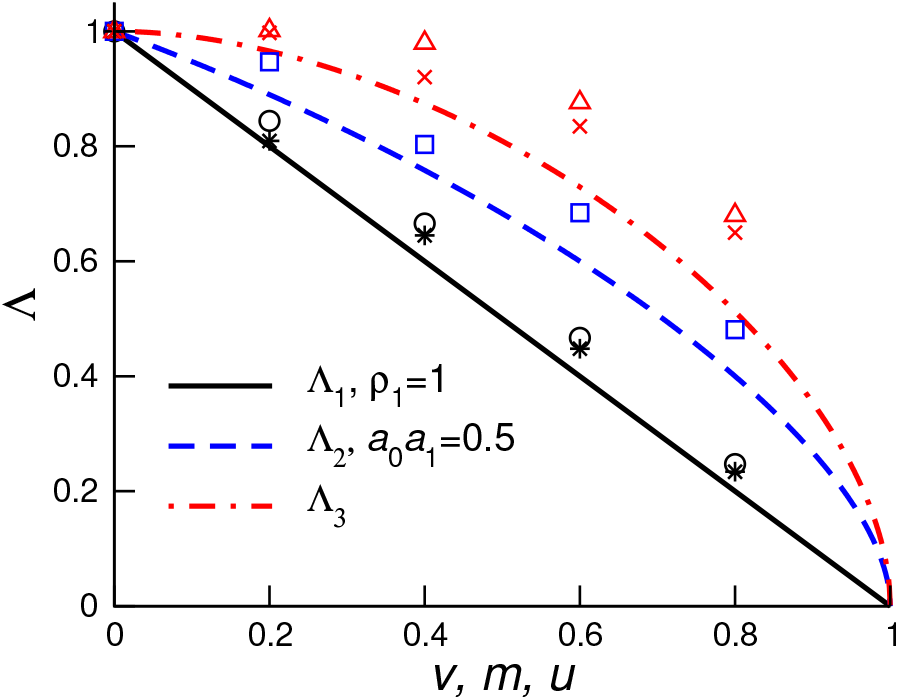
Largest eigenvalue as a function of the relevant parameter for the listed containment strategies. Lines are the analytical predictions (20) and the symbols numerical estimates: circles for vaccinated fraction *v*, squares for mask wearing fraction *m*, triangles for app user fraction *u*, * for vaccinated fraction *v* together with *m* = 0.2, and Π for app user fraction *u* together with *m* = 0.2.

Now I combine the containment strategies. Substituting equations (12)-(19) into equation (11) we obtain

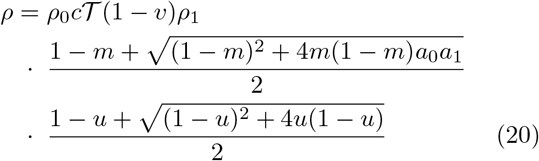

where *ρ*_0_ = ⟨*β*^2^ ⟩*/*⟨ *β* ⟩*γ* is the basic reproductive number of the standard SIR model. This equation is the starting point for a comprehensive understanding of how intervention strategies impact the expected reproductive number. In the absence of contact heterogeneity (⟨*β*^2^⟩ =⟨ *β* ⟩^2^) and no interventions (*c* = 1, 𝒯 = 1, *v* = 0, *ρ*_1_ = 1, *m* = 0, *u* = 0), we recover *ρ* = *ρ*_0_. In the present of multiple containment strategies, we can use (20) to estimate the aggregate impact. For example, combining a 50% of mask users with a 50% of smartphone tracing app users will reduce the reproductive number by about a half. Add to that a 50% vaccination and it will reduce the reproductive number by about a third.

I have performed agent based simulations to test Eq. (20). I have generated a virtual city where places are represented by nodes in a network and the flow of people between two places is represented by a link between the associated nodes [11]. Agents are located at different places and they switch place following the network links, at certain rate that varies between individuals. A SIR model is simulated in the virtual city introducing a patient zero and constraining the disease transmission to individuals at the same place [11]. The value of Λ is estimated as Λ(*x*) = *ρ*(*x*)*/ρ*(0), where *x* = *v, m* or *u* and *ρ*(*x*) is obtained from a fit of (6) to the early growth phase of the numerical data [11]. The analytical prediction (20) is in good agreement with the numerical estimates 2, although the theoretical line always underestimates the numerical values. The underestimation can be due to the contribution of eigenvalues besides the largest.

In conclusion, the analytical predictions estimate and explain the impact of multiple containment strategies on the reproductive number of an epidemic outbreak.

## Data Availability

NA

## Notes

### Competing Interest Statement

The authors have declared no competing interest.

### Summary of Updates

This new version adds numerical simulations to text the theoretical predicions.

