## Supplementary material for "Multi-type branching and graph product theory of infectious disease outbreaks"

Alexei Vazquez[1,*]

[1]*German Aerospace Center (DLR), Institute for the Protection of Terrestrial Infrastructures, Rathausallee 12, 53757 Sankt Augustin, Germany*

(Dated: January 22, 2021)

*

### MAPPING TO COMPARTMENT MODELS

In the compartment SIR model with $N$ agents one focuses on the total number of susceptible ($S$), infected ($I$) and removed ($M$) agents, with $S+I+M=N$. In the multi-type version $S$, $I$ and $M$ are vectors, with components counting the individuals according to type. The transitions between these different compartments is dictated by the system of first order differential equations

$$\begin{aligned}\frac{1}{\gamma}\dot{I} &= IR\frac{S}{N} - I\\ \frac{1}{\gamma}\dot{M} &= I\\ S &= N - I - M\end{aligned} \tag{1}$$

where $R$ is the reproductive number matrix and $\gamma$ is the recovery rate. I note that $R$ is proportional to the ratio between the transmission and recovery rates. Therefore, $\gamma R$ is proportional to the disease transmission rate. In the case where $R$ is diagonalized by a transformation $P$, we can multiply from the left by $P$ and from the right by $P^{-1}$ to obtain

$$\begin{aligned}\frac{1}{\gamma}\dot{x} &= xR_D(I_N - x - y) - x\\ \frac{1}{\gamma}\dot{y} &= x\end{aligned} \tag{2}$$

where $I_N$ is the identity matrix of size $N$, $R_D$ is the diagonal form of $R$ and

$$\begin{aligned}x &= PIP^{-1}/N\\ y &= PMP^{-1}/N\\ R_D &= PRP^{-1}\end{aligned} \tag{3}$$

Equation (2) represents the dynamics of the eigenmodes of the disease spreading process in the multi-type population. When $x, y \ll 1$ then (2) can be integrated analytically obtaining

$$x_i(t) \sim e^{(\lambda_i - 1)\gamma t} \tag{4}$$

where $\lambda_i$ are the eigenvalues of $R$. When the largest eigenvalue of $R$, $\rho = \max\{\lambda\}$, satisfies $\rho > 1$, then the associated eigenmode will increase exponentially in time, as deduced from the branching process formulation as well. More generally, equation (2) can be integrated to obtain the time-dependent evolution.

### AGENT BASED SIMULATIONS OF EPIDEMIC OUTBREAKS

#### Barabási-Albert city

I have developed an agent based model inspired in the human mobility pattern of Portland [1]. The Portland study simulated the movements of about 1000000 individuals between about 100000 places within the city of Portland. The study reported that the distribution of the number of places visited by an individual has a maximum at 2 places and an exponential tail. In contrast the number of visitors per day at a given place follows a broad distribution with a power law tail, with exponent $-2.8$. Using that study as a blueprint I have developed and agent based simulation of people mobility within a city.

Given that the places have a broad distribution of visitors per day with a power law exponent close to -3, it is natural to think about places as nodes on a Barabásis-Albert graph. If places are nodes in a graph and agents switch place following the graph edges, at an average switching rate $\langle\omega\rangle$, then the steady state balance of outgoing and incoming visitors to a place is given by

$$n_i\langle\omega\rangle = k_i\langle\omega\rangle\langle n_i\rangle \tag{5}$$

where $k_i$ is the degree of place $i$ in the place-to-place graph. This detailed balance warranties that the number of visitors to a place is proportional to the degree of the place in the place-to-place graph. Since the latter is a Barabási-Albert network this warranties that the distribution of the number of visitors to a place follows a power law distribution with exponent -3, close to what observed for the city of Portland.

In turn, I will assume that individuals switch location at a rate $\omega_i$, where the $\omega_i$ are random variables with a gamma distribution

$$\mathrm{Prob}(\omega_i = \omega) = \frac{1}{\omega^*}\left(\frac{\omega}{\omega^*}\right)^{s-1} e^{-\omega/\omega^*} \tag{6}$$

I will set $\omega^* = 1$ switch per day and $s = 2$, which gives a mode at 1 switch per day. This means that individuals will be in about two places per day, one where they started the day and the other where they switch to, as observed for the Portland city simulation.

The place-to-place graph introduced above does not represent connectivity by geographic distance. It is a graph representing the flow of people between places. Furthermore, this is an annealed city, in the sense that individuals are not assigned to quenched locations to where they always comeback.

#### Epidemic outbreaks

To simulate epidemic outbreaks I will use the susceptible, infected and removed (SIR) compartment model on a Barabaásis-Albert city. Individuals can be in the states susceptible, infected and removed. Infected individuals get in contact with other individuals within the same location at a rate $\xi$. If the other individual is in the susceptible state then it is switched to the infected state. Infected individuals are removed at a rate $\gamma$, to model death, hospitalization or any other process that removes infected individuals from the disease transmission chain. To run the infectious model I will use parameters close to the SARS-CoV-2 virus. The infectious periods is about 3 days. Thus I set $\gamma = 1/3$ per day. The value of $\xi$ will be set high enough that we obtain an exponential growth for the non-pharmaceutical interventions investigated. This is done such that we can obtain an estimate of the outbreak growth rate, which can be related to the largest eigenvalue of the reproductive number matrix. Thus, I set $\xi = 4$ attempts of transmissions per day.

The full stepwise procedure to simulate epidemic outbreaks is

1. Set the number of agents to $n_a = 1000000$ and the number places $n_p = 100000$.

2. Generate a Barabási-Albert model with $m = 2$. Specifically, start with a complete graph of $m + 1$ nodes. Then add new nodes one at the time up to $n_p$ nodes. Each time a node is added, it is connected to $m$ nodes in the pre-existing graph. The node to which each of the $m$ edges is attached to is selected with a probability proportional to the nodes degree.

3. Assign switching rates $\omega_i$ to each $i = 1, \ldots, n_a$ agent extracted from the gamma distribution (6).

4. Assign each agent to a place with a probability proportional to the place degree in the Barabási-Albert network.

5. Run the city simulation for $10n_a$ switching events to randomize the distribution of agents across places. At each step, select an agent with a probability proportional to the agent switching rate $\omega_i$. Then select a place at random from the neighbours of the current agent place in the Barabási-Albert graph. Move the agent to that new place.

6. Overlay the epidemic outbreak on the city simulation. Set time to $t = 0$, set one individual, sampled uniforminly from all individuals, to the infected state, and set all other individuals in the susceptible state. At each step select an event type among switching place, disease transmission and recovery according to the rates

   - Switching rate $\sum_i \omega_i$: Select an agent with a probability proportional to the agent switching rate $\omega_i$. Then select a place at random from the neighbours of the current agent place in the Barabási-Albert graph. Move the agent to that new place.
   - Transmission rate $\xi n_I$: Select an infected agent sampled uniformingly from all infected agents (the source of transmission). Select an agent from the place where the selected infected agent is located (the target of transmission). If that agent is in the susceptible state switch the agent to the infected state. Otherwise do nothing.
   - Removing rate $\gamma n_I$: Select an infected agent sampled uniformingly from all infected agents. Set the agent's state to removed.

where $n_I$ is the number of current individuals in the infected state. Update time $t \to t + \Delta t$, where $\Delta t$ is extracted from an exponential distribution $\lambda_{Total} e^{-\lambda_{Total} \Delta t}$, where $\lambda_{Total} = \sum_i \omega_i + \xi n_I + \gamma n_I$.

#### Interventions

The interventions were implemented as follows

- Vaccination: Individuals are categorized as vaccinated or not with probability $v$. The transmission rule was modified to include the constraint that transmission will not happen if the target is vaccinated.
- Mask use: Individuals are categorized as wearing a mask or not with probability $m$. The transmission rule was modified to include the constraint that if the source and the target wear mask transmission will not happen, if the source wears a mask but the target does not then transmission happens with probability $a_0$, and if the source does not wears a mask but the target does then transmission happens with probability $a_1$. Here I will use $a_0 = a_1 = \sqrt{0.5}$ for the purpose of illustration.
- Phone App: Individuals are categorized as downloading the phone tracing app or not with probability $u$. The removing rule was modified to include the contact tracing based on the phone tracing app. For the sake of simplicity I have assumed that the tracing is triggered at the point when an infectious agent is removed (the tracing seed), which is when the agent should manifest symptoms and therefore being detected. Then I implement backward and forward tracing starting from the seed agent, constrained to agents that carry the phone tracing app. In the tracing, I recursively identify the agent that transmitted/received the disease to/from the seed agent and keep doing that backward/forwards in the disease transmission chain, constrained to being app users. All the traced agents are set to the removed state.

### ESTIMATION OF $\Lambda$

To estimate $\Lambda$ we calculated the number of infected individuals $n(t) = \dot{I}$ as a function of time $(t)$ average over 100 simulated cities and then outbreaks and binned in steps of 1 day. $n(t)$ was fit from $t = 0$ to $t = 0.8 t_0$ to the exponential growth $n(t) = n_0 e^{\mu t}$, where $t_0$ is the time where $n(t)$ is maximum.. The value of $\rho$ was calculated taking into account that $n(t) \sim e^{(\rho - 1)\gamma t}$, i.e.

$$\rho = \frac{\mu}{\gamma} - 1 \tag{7}$$

For a given intervention parametrized by $x$, $\Lambda(x)$ was calculated as

$$\Lambda(x) = \frac{\rho(x)}{\rho(0)} \tag{8}$$

### RESULTS

The number of individuals at a given place (place size) scales linearly with the degree and the distribution of place sizes has a power law tail with a power law exponent close to -3. Data obtained after running one realization of steps 1-5 above. Left linear binning. Right logarithmic binning.

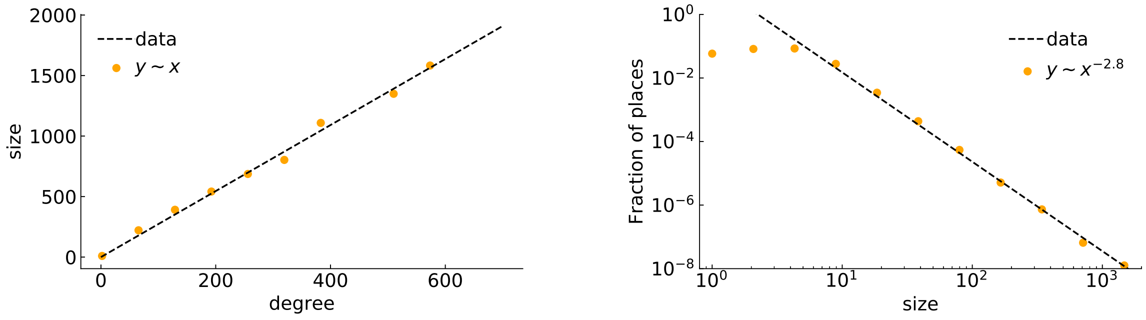


Number of infected individuals and estimated $\Lambda$ vs $v$ for $m = u = 0$.

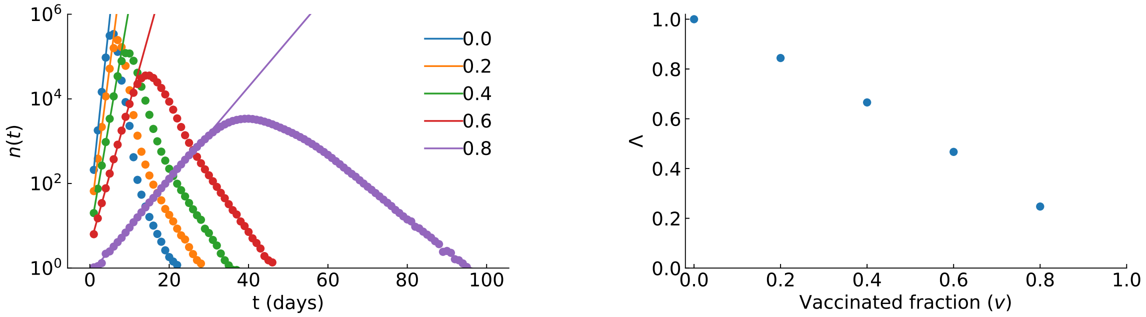


Number of infected individuals and estimated $\Lambda$ vs $v$ for $m = 0.2$ and $u = 0$.

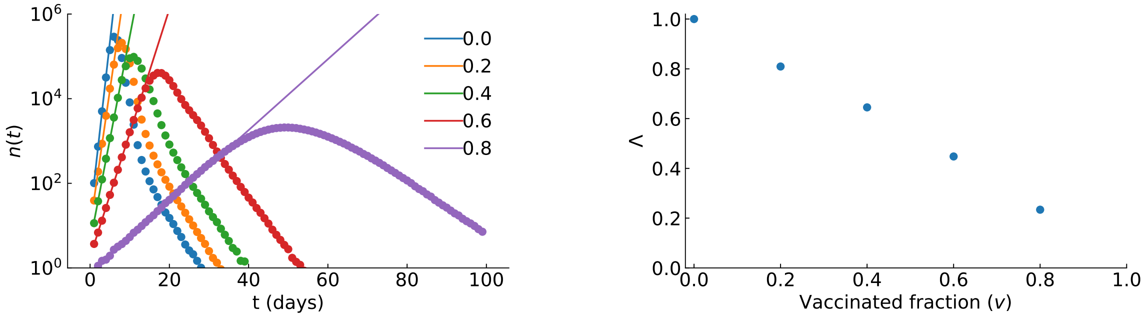


Number of infected individuals and estimated $\Lambda$ vs $m$ for $v = u = 0$.

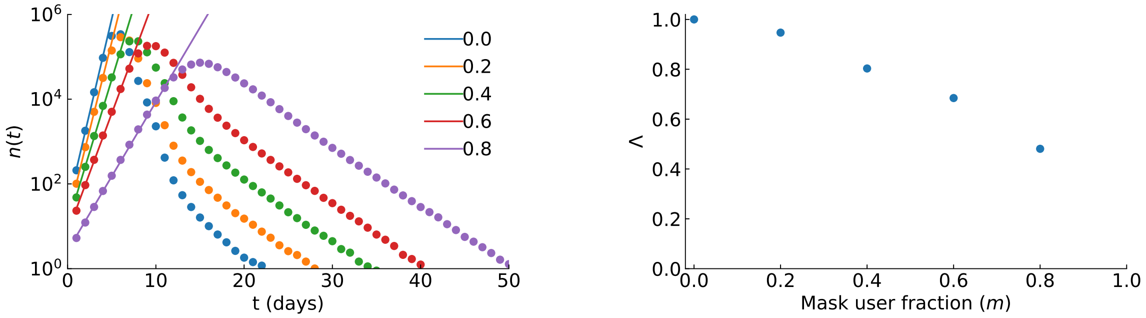

Number of infected individuals and estimated $\Lambda$ vs $u$ for $v = m = 0$.

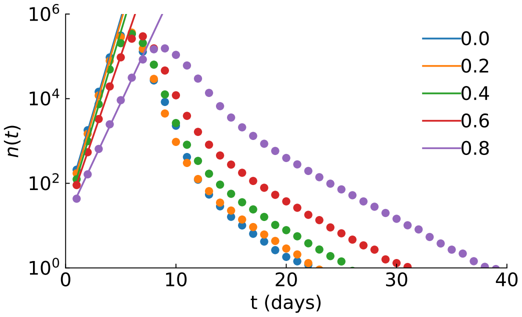


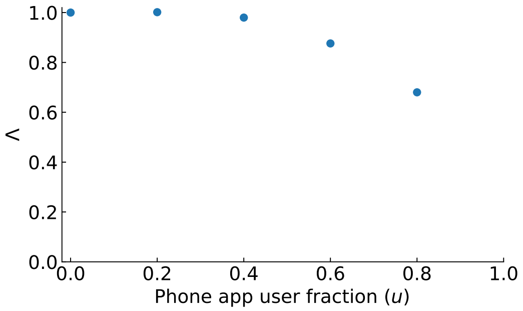


Number of infected individuals and estimated $\Lambda$ vs $u$ for $m = 2$ and $v = 0$.

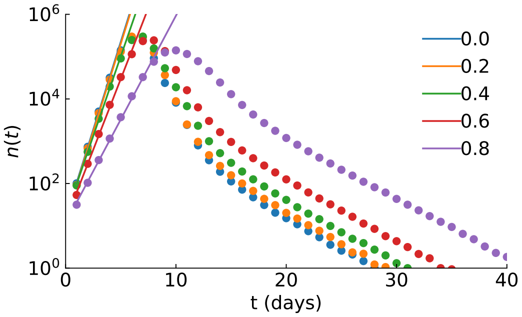


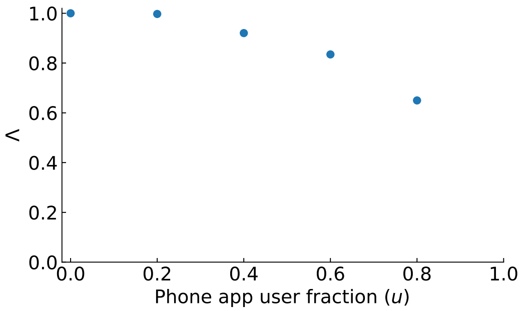
